# Epidemiology and Risk Factors of Cutaneous Leishmaniasis in the West Bank, Palestine: A Cross-Sectional Study

**DOI:** 10.1101/2024.11.26.24318023

**Authors:** Elaf AbuZaaroor, Kamal AlShakhra, Moath Alawneh

## Abstract

**Background:** Leishmaniasis Is a vector-borne disease caused by obligate intracellular protozoan flagellate parasite, that is transmitted by the bite of infected female phlebotomine sandflies. Cutaneous Leishmaniasis (CL) is endemic in the Middle East, it’s a major public health concern in Palestine, the current control strategies for Leishmaniasis depend on reservoir and vector control, active case detection and treatment of their disease and the use of insecticides.

**Objectives:** This study aims to describe the epidemiology, clinical features, and risk factors of CL in the West Bank, Palestine, from 2020 to 2023.

**Methods:** A retrospective study included all cases of CL that had been reported to the Leishmaniasis Surveillance System in the Department of preventive medicine at the Palestinian Ministry of Health during the period from 2020 to 2023. A total of 322 cases were reported and met the case definition. demographic details, lesion characteristics, and environmental risk factors., Independent t-test and ANOVA were used to obtain the differences between the groups according to lesion duration, number of lesion.

**Findings:** A total of 322 cases of Cutaneous Leishmaniasis were diagnosed in preventive medicine departments in West bank during the period 2020 till 2023, the ages of patients ranges between less than one year to 82 years old, most cases were young, with mean age of 24 years old, the male to female ratio was 3:2, most patients were illiterate,.

The number of skin lesions was significantly higher in males, farmers and most patients have face lesions mainly nodules. while the duration of the skin lesions was significantly higher in older patients.

**Conclusions:** CL is endemic in the West Bank. Public health strategies should emphasize early detection, community awareness, and targeted vector control measures.

**Funding:** No funding was provided for this research.

## 1. Introduction

### 1.1 Background

Leishmaniasis is a vector-borne disease that is transmitted via female phlebotomine sandflies and caused by obligate intracellular protozoa called *Leishmania*, there are 30 Leishmania species that infect mammal; out of these 21 are known to infect humans ^(1)^. It is endemic in 98 countries and 3 continents ^(1)^. Leishmaniasis is subdivided into 3 types: cutaneous, mucocutaneus, and visceral which is the most serious form, also known as (Kala Azar). Cutaneous leishmaniasis is the most common type, and almost 95% of its cases are found in the Mediterranean region, the Middle East, North and South America, and Central Asia ^(2)^.

Skin manifestation of Cutaneous Leishmaniasis, are mostly ulcers, that leave permanent scars and cause disability ^(2).^Diagnosis of leishmaniasis is based on endemicity, clinical symptoms, and laboratory tests.

Leishmaniasis is a significant, neglected, zoonotic disease and a great challenge to public health, especially in poor and underdeveloped countries ^(3)^. Leishmaniasis, is the third most important arthropod-borne diseases in terms of the global burden of diseases ^(4)^. That poverty, malnutrition, immigration, overcrowding, deforestation, urbanization and new settlements, play a great role in increasing the incidence of CL ^(2)^. Up to date there are no available drugs or vaccines to prevent infections.

### 1.2. Epidemiology of Leishmaniasis

Leishmaniasis is endemic in approximately 90 countries in the tropics, subtropics, and southern Europe. The ecologic settings range from rain forests to deserts. Leishmaniasis usually is more common in rural than in urban areas, but it is found in the peripheries of some cities. Climate and other environmental changes attribute to expand the geographic range of the sand fly vectors and the areas in the world where Leishmaniasis is found ^(1)^. It has certainly a wider geographical distribution than before; it is now reported in areas that formerly were non-endemic, and more than 60% of the new cases are found in 6 countries: Algeria, Afghanistan, Brazil, Colombia, Islamic Republic of Iran, Pakistan, Peru, Saudi Arabia, and Syria ^(2).^

An estimate of around 0.7 to 1 million new cases of Leishmaniasis are reported annually ^(2)^ .is found in people on all continents except Australia and Antarctica, cases are reported in:

- In the Eastern Hemisphere (the Old World), Leishmaniasis is found in some parts of Africa (particularly in the tropical region and North Africa), Asia, the Middle East, and southern Europe ^(1)^.
- In the Western Hemisphere (the New World), it is found in some areas of Mexico, Central America, and South America. But not in Chile or Uruguay. Occasional cases of cutaneous Leishmaniasis have been reported in Texas and Oklahoma ^(1)^.

### 1.3. Leishmaniasis in Palestine

Despite the significant breakthroughs in reducing the disease burden, Leishmaniasis remains a significant public health challenge in Palestine. Like all neighboring countries it is a reportable infection; Jordan, Syria and Saudi Arabia. Notification of the Ministry of Health (MOH) of Leishmaniasis in Palestine is required by law.

Ministries of Health collect data for therapeutic and control strategies. It is reported in all Palestinian districts except Gaza strip. North of Palestine is the center of a region where simple CL is hyper endemic ^(5)^.These villages have closely related and characteristic topographies, ecology, and cultivation that undoubtedly impact Leishmania incidence and vector populations. Besides, these are areas with a relatively low socioeconomic level, poor health services, many uncontrolled dumping sites, and an abundance of domestic animals such as chicken and goats. These conditions have been identified as risk factors of human Leishmaniasis ^(6)^.

According to world health organization there is little information about the incidence, prevalence or distribution of human disease in occupied Palestinian territory, although there have been detailed studies on the vectors and reservoir hosts of zoonotic cutaneous Leishmaniasis on the West Bank ^(7)^.

### 1.4. Major risk factors for Leishmaniasis

Leishmaniasis is caused by more than 20 species of Leishmania parasites that are spread by about 30 types of phlebotomine sand flies; particular sand flies spread particular species of the parasite. The sand fly vectors generally are the most active during twilight, evening, and night-time hours (1).

#### 1.4.1. Socioeconomic conditions

Individuals of all ages who have not previously had the disease are at risk for Cutaneous Leishmaniasis infection, if they are exposed to sand fly bites ^(8)^, but according to the previous studies and available data most cases are young people ^(9) (10)^. The high prevalence rate in children is attributed to their habits ^(8)^.

Poverty increases the risk for Leishmaniasis. Domestic sanitary conditions (lack of waste management or open sewerage) and poor housing may increase sandfly resting sites and breeding, in addition to their access to humans. Sandflies are attracted to crowded housing. Human behavior, such as sleeping outside or on the ground make it is easier for sandflies to bite them and feed on their blood ^(2)^.

Eid et al found that most infected men were working in agriculture while most women were housewives, besides the educational level was 6.2 years on average ^(9)^. Another supporting study found that the majority of CL cases were men ^(9) (10)^, farmers with low schooling, and residents in rural areas^(10)^.

Another significant risk factor is the presence of domestic animals such as dogs, pigs, or chickens and raising livestock on the spot, that provides a favorable environment to the biological development of sand flies ^(11)^, and suggested that animals could attract the vectors closer to humans ^(12) (13)^. Most of the cases raised hens (86.5%), while less than half had dogs (40.4%) ^(9)^. On the other hand other studies have found a protective role in animals presence arguing that animals would be a preferred source of blood for vectors ^(14)(15)^.

Housing conditions of the cases were studied and it was found that if walls, roofs, and floors are not made of tough materials, cracks may be formed, becoming a gateway for vectors into the houses ^(9)^. another important factor is the increasing density of the population and its relationship with making of the favorable environments to the multiplication of the phlebotomy vector ^(11)^.

The incidence of Leishmaniasis is usually affected by changes in urbanization, deforestation, or the human movement into forested areas. Climate change affects the spread of Leishmaniasis though changes in temperature and rainfall, which affect the geographic distribution and size of sand fly populations. Drought, flood and famine also cause migration of people into areas where the transmission of the parasite is high ^(2)^.Infection mainly occurs during the summer months, which causes the disease to appear later during the winter months ^(16)^.

### 1.5. Clinical manifestation

Cutaneous lesions can either be a single, limited skin lesion or, large, multiple locally destructive skin lesions, which appear after an incubation period, that can be many months ^(17)^. Spontaneous healing usually results in lifelong immunity ^(18)^.

CL lesions usually appear on body parts that are exposed to the environment, such as the face, forearms, and lower legs. The infection generally presents as a painless, brownish, erythematous papule after a long (2-8 months) incubation period. These papules then gradually enlarge and turning into a nodule or plaque within 6 months ^(16)^.

### Aim of the study

This study seeks to bridge the knowledge gap by describing the epidemiology, clinical features, and environmental risk factors of CL in Palestine, offering insights into public health interventions.

## Methods

### Study design

This is a retrospective study included all cases of cutaneous Leishmaniasis which had been reported to the Leishmaniasis Surveillance System in the preventive medicine Department at the Palestinian Ministry of Health from January 2020 to December 2023. A total of 322 cases were reported and met the case definition.

### Data collection

The available and necessary data were retrieved from the surveillance system. Data included patient’s age, gender, residency, occupation, reporting date, number, site, form of the lesions, and environmental risk factors.

### Data analysis

Statistical analysis was performed using SPSS, version 21.0 (SPSS Inc., Chicago, IL, USA). Continuous variables are expressed as the means ± standard deviations (SD) and categorical variables as frequencies and percentages. Independent t-test and Chi-square test (or Fisher test) were used to compare means and proportions between groups. Multivariate Binary logistic regression was used to assess the association between different factors and number of lesions. The level of significance was set at 5%.

### Ethics Approval

Ethical approval was obtained from the Palestinian Ministry of Health. Persons were anonmyized.

## Results

322 patients were diagnosed to have cutaneous Leishmaniasis in 2020 till 2023 in the west bank, the ages of patients ranges between less than one year to 82 years old, with mean 24 years old, and median is 17 years old and Mode 11years old, the distribution of the cases by gender and age group is expressed in figure 1.

**Figure 1.**
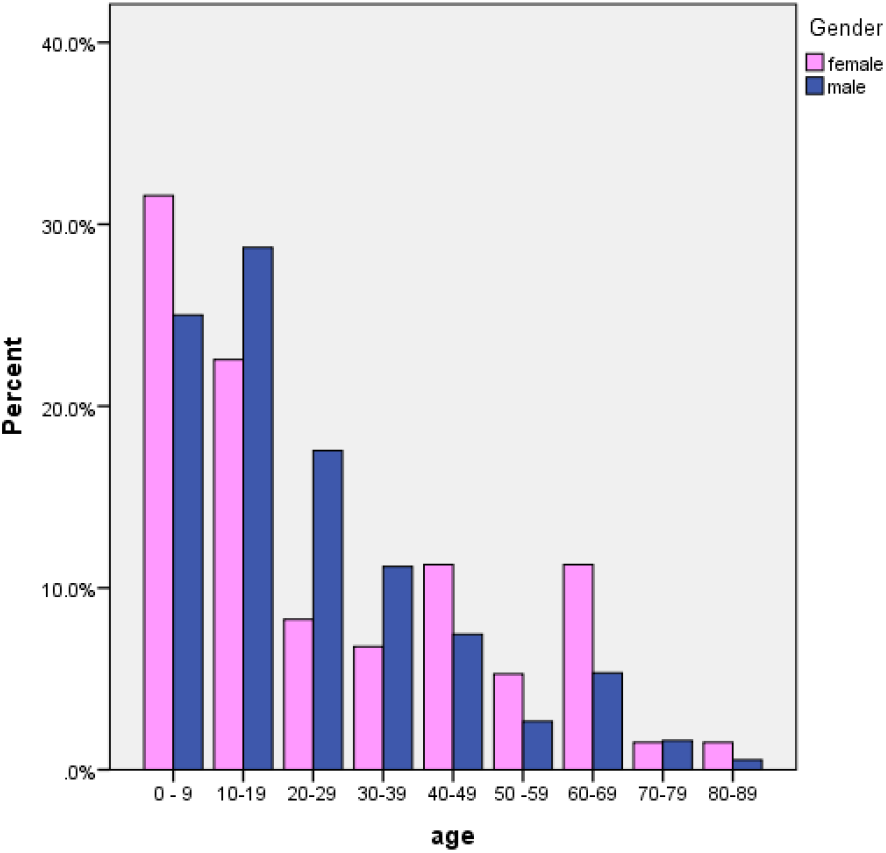
Distribution of cases by gender and age groups.

58.6% of the cases were males, while 41.4 % were females, the male to female ratio was 3:2; the mean of the ages were 22 and 26 years old respectively. This difference in age was insignificant (p value = 0.109).

Out of the 322 cases, most were from Tubas 19.9%, then Hebron 17.7%and Jericho 13.7%. Most cases were students (34.2 %), workers 19.4%), housewives and children (14.8 and 15.2% respectively).

The average years of education for the Patient was 4.4 years old, fathers of the patients 4.2, while mother’s years of education was 3.8.

Regarding the presence of animals, the percentages of patients who had confirmed to have animals were :Domestic dogs 54%, Stray dogs 56.5%, Stone fences 54%, Hens 47%, Foxes jackals 32%, Rocky hyrax 42.2%, Rats 39.1% And Domestic animals 57.5%

53% of the cases had caves around their houses; nearly 65% of the cases have more than five family members inside the home.

Most of the patients had single skin lesion at presentation, while 39.5 % had multiple, of these 22.5% had 3 or more skin lesions, most of the skin lesions in the patients were on their faces; 57.1% of the patients had face lesions, 48.4% of the cases have lesions over their limbs. And about 52% had nodules, 41% were presented with skin ulcers.

9% of the patients were diagnosed based on clinical basis and epidemiological data only without further laboratory investigations, while 91% of the cases were confirmed by laboratory tests, (mainly direct skin smear of lesion (90.7%).

About one half of the cases were diagnosed in 2020, during winter and early spring (January till May), (figure 2) the number then decreases and start to rise again in October, the average duration of the lesion at presentation is about 3 months (as shown in Figure 3).

**Figure 2.**
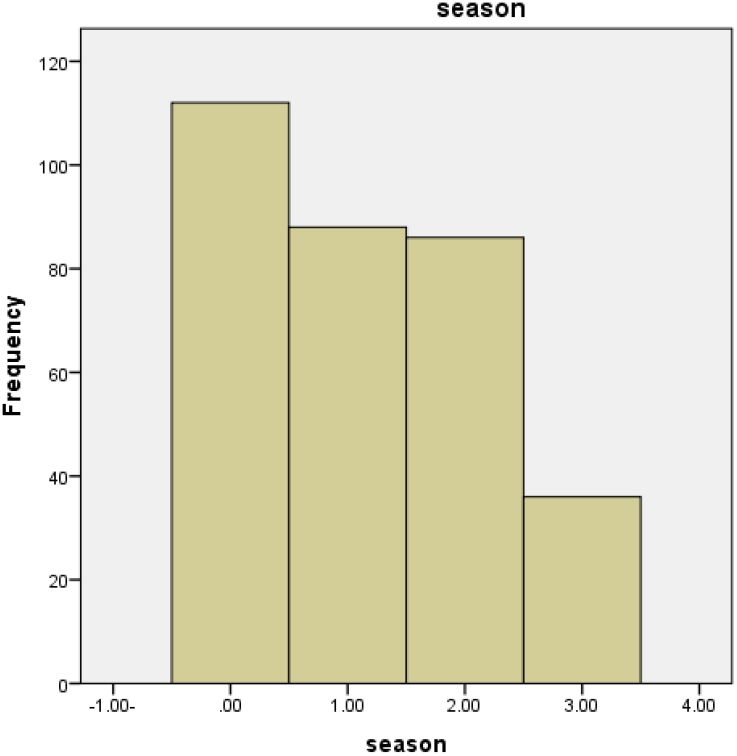
Distribution of cases in different seasons.

**Figure 3.**
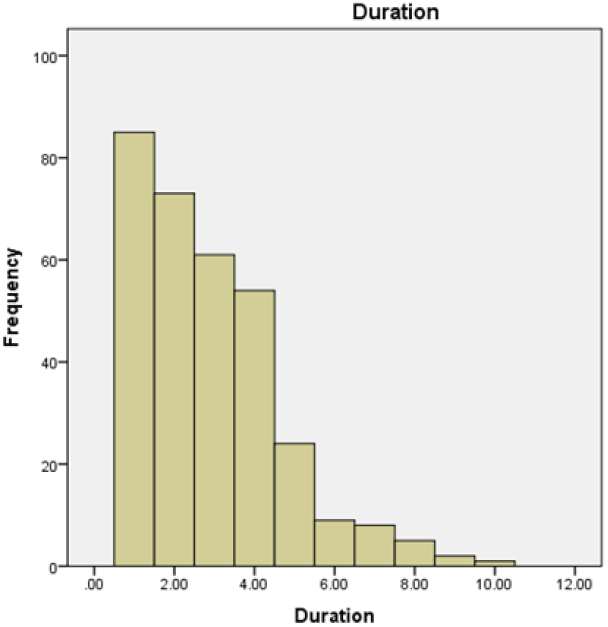
Distribution of the duration of skin lesions.

**Figure 4.**
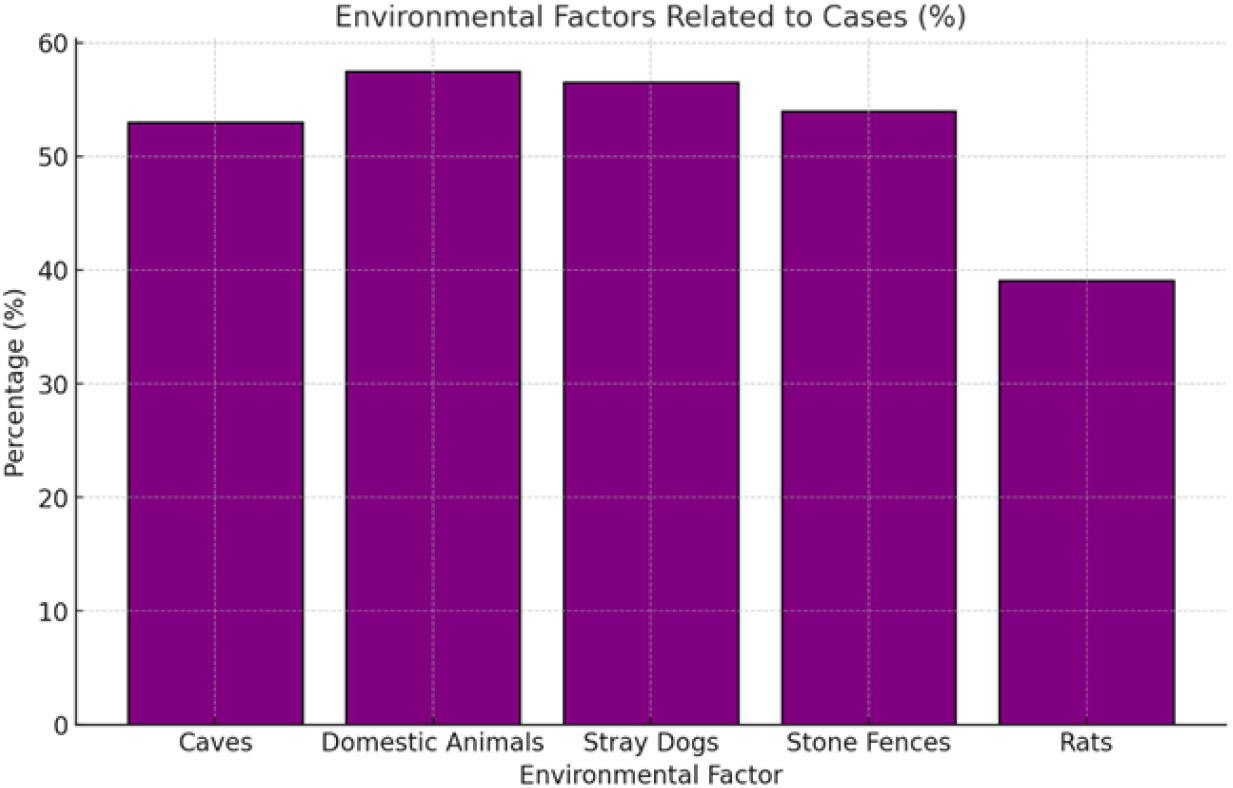
Distribution of the environmental factors related to cases.

The association between the demographic risk factors with the number and the duration of the lesions is summarized in Table 1. The association between the environmental risk factors and the number and the duration of the lesions is summarized in Table 2.

**Table 1:**
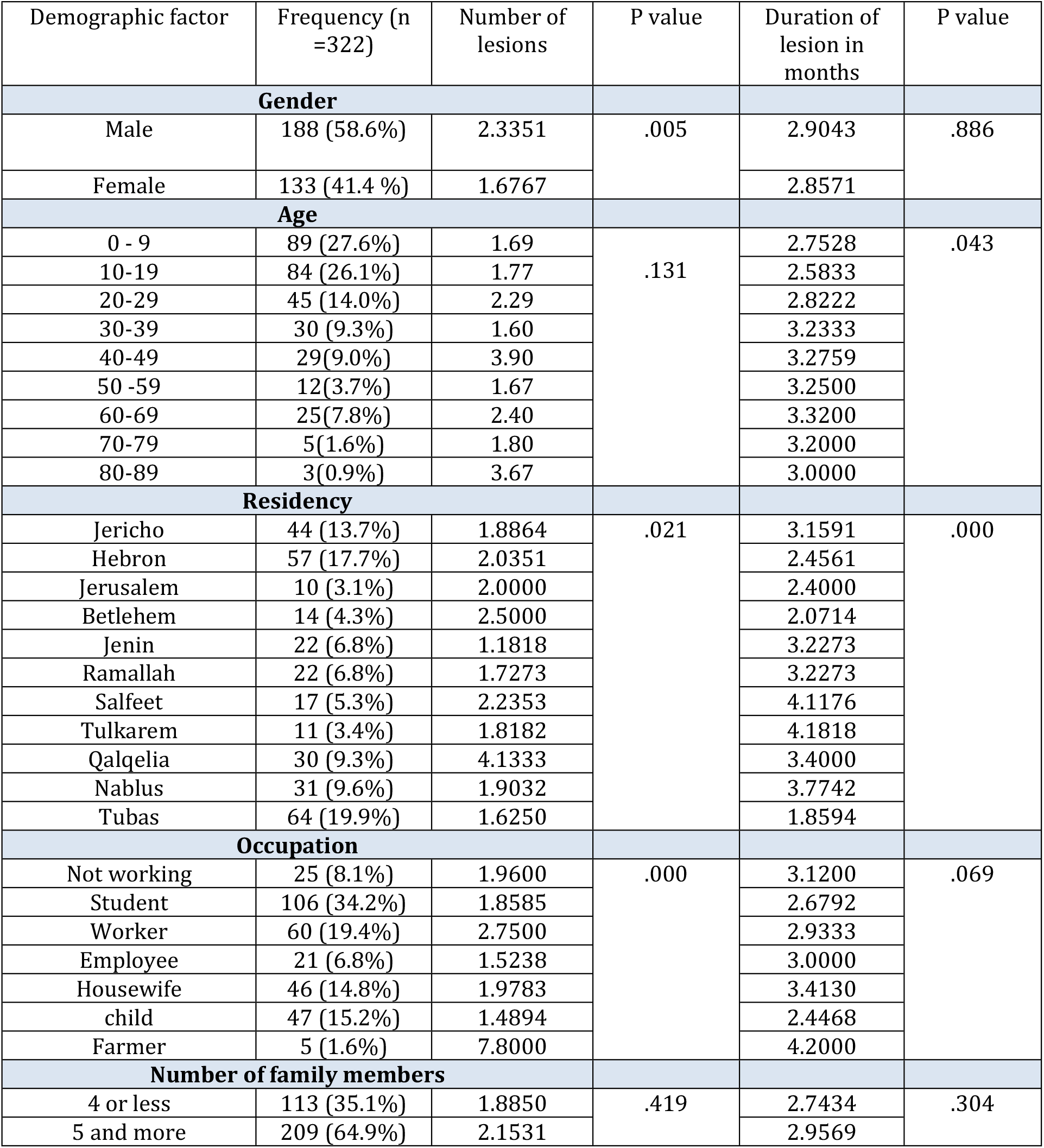
The demographic distributions of leishmaniasis cases.

| Demographic factor | Frequency (n =322) | Number of lesions | P value | Duration of lesion in months | P value |
| --- | --- | --- | --- | --- | --- |
| **Gender** |  |  |  |  |  |
| Male | 188 (58.6%) | 2.3351 | .005 | 2.9043 | .886 |
| Female | 133 (41.4 %) | 1.6767 |  | 2.8571 |  |
| **Age** |  |  |  |  |  |
| 0 - 9 | 89 (27.6%) | 1.69 | .131 | 2.7528 | .043 |
| 10-19 | 84 (26.1%) | 1.77 |  | 2.5833 |  |
| 20-29 | 45 (14.0%) | 2.29 |  | 2.8222 |  |
| 30-39 | 30 (9.3%) | 1.60 |  | 3.2333 |  |
| 40-49 | 29(9.0%) | 3.90 |  | 3.2759 |  |
| 50 -59 | 12(3.7%) | 1.67 |  | 3.2500 |  |
| 60-69 | 25(7.8%) | 2.40 |  | 3.3200 |  |
| 70-79 | 5(1.6%) | 1.80 |  | 3.2000 |  |
| 80-89 | 3(0.9%) | 3.67 |  | 3.0000 |  |
| **Residency** |  |  |  |  |  |
| Jericho | 44 (13.7%) | 1.8864 | .021 | 3.1591 | .000 |
| Hebron | 57 (17.7%) | 2.0351 |  | 2.4561 |  |
| Jerusalem | 10 (3.1%) | 2.0000 |  | 2.4000 |  |
| Betlehem | 14 (4.3%) | 2.5000 |  | 2.0714 |  |
| Jenin | 22 (6.8%) | 1.1818 |  | 3.2273 |  |
| Ramallah | 22 (6.8%) | 1.7273 |  | 3.2273 |  |
| Salfeet | 17 (5.3%) | 2.2353 |  | 4.1176 |  |
| Tulkarem | 11 (3.4%) | 1.8182 |  | 4.1818 |  |
| Qalqelia | 30 (9.3%) | 4.1333 |  | 3.4000 |  |
| Nablus | 31 (9.6%) | 1.9032 |  | 3.7742 |  |
| Tubas | 64 (19.9%) | 1.6250 |  | 1.8594 |  |
| **Occupation** |  |  |  |  |  |
| Not working | 25 (8.1%) | 1.9600 | .000 | 3.1200 | .069 |
| Student | 106 (34.2%) | 1.8585 |  | 2.6792 |  |
| Worker | 60 (19.4%) | 2.7500 |  | 2.9333 |  |
| Employee | 21 (6.8%) | 1.5238 |  | 3.0000 |  |
| Housewife | 46 (14.8%) | 1.9783 |  | 3.4130 |  |
| child | 47 (15.2%) | 1.4894 |  | 2.4468 |  |
| Farmer | 5 (1.6%) | 7.8000 |  | 4.2000 |  |
| **Number of family members** |  |  |  |  |  |
| 4 or less | 113 (35.1%) | 1.8850 | .419 | 2.7434 | .304 |
| 5 and more | 209 (64.9%) | 2.1531 |  | 2.9569 |  |

**Table 2:** The association between the environmental risk factors and the number and the duration of the lesions.

|  | Frequency (n =322) | Number of lesions | P value | Duration | P value |
| --- | --- | --- | --- | --- | --- |
| **Domestic dogs** |  |  |  |  |  |
| No domestic dogs | 148 | 1.9797 | .645 | 3.2162 | .002 |
| Domestic dogs | 174<br>54% | 2.1264 |  | 2.5977 |  |
| **Caves and crevices** |  |  |  |  |  |
| No caves and crevices | 152 | 2.5132 | .007 | 3.1118 | .030 |
| Caves and crevices | 169<br>53% | 1.6568 |  | 2.6805 |  |
| **Stray dogs** |  |  |  |  |  |
| No Stray dogs | 140 | 2.0929 | .852 | 2.9857 | .359 |
| Stray dogs | 182<br>56.5% | 2.0330 |  | 2.8022 |  |
| **Stone Fences** |  |  |  |  |  |
| No Stone Fences | 147 | 2.0952 | .834 | 3.0204 | .201 |
| Stone Fences | 175<br>54% | 2.0286 |  | 2.7657 |  |
| **Hens** |  |  |  |  |  |
| No Hens | 171 | 2.2632 | .170 | 3.0058 | .184 |
| Hens | 151<br>47% | 1.8278 |  | 2.7417 |  |
| **Foxes and Jukes** |  |  |  |  |  |
| No Foxes and Jukes | 219 | 2.1598 | .354 | 2.9635 | .231 |
| Foxes and Jukes | 103<br>32% | 1.8447 |  | 2.7087 |  |
| **Rocky Hyrax** |  |  |  |  |  |
| No RockyHyrax | 186 | 2.0161 | .752 | 3.0753 | .022 |
| RockyHyrax | 136<br>42.2% | 2.1176 |  | 2.6176 |  |
| **Rats** |  |  |  |  |  |
| No Rats | 196 | 2.2449 | .143 | 2.9235 | .602 |
| Rats | 126<br>39.1% | 1.7698 |  | 2.8175 |  |

## Discussion

The mean age of our study participants is about 24 years old. More than 50% of patients were below 19 years old. This means that Children are the most affected age group. Our findings are similar to previous studies in Palestine ^(19, 20)^. The increase in infection prevalence in children may be due to lower exposure to infectious sand-fly bites in young children, but likely due to the acquired immunity to the parasite increases with age.

However, poor standardization of serological tests makes it difficult to compare data from various studies and determine the differences in observed age patterns ^(21).^

In the current study, among various occupations, the highest prevalence of leishmaniasis was found in school and university students. Most students aged 4–23 year old and have not been infected by the disease; they comprise a high proportion of patients, the same finding of other studies ^(22)^.

Most cases were males, with a proportion of 3 to 2; this is similar to other previous studies worldwide; biological sex-related differences may play a role in the pathogenesis of this disease^(23)^.

More than 70% of the patients have more than 5 family members in the house; this can be explained by the fact that sandflies are more attracted to crowded housing as these provide a good source of blood-meals ^(2)^. Poverty increases the risk for Leishmaniasis. Poor sanitary conditions (like lack of waste management or open sewerage) may increase sand-fly breeding and resting sites, as well as their access to humans ^(2)^.

More than 60% of the cases reported that they had domestic animals, mainly dogs, previous case control study in Nepal had reported that ownership of animals and keeping animals inside the house increases the risk of Leishmaniasis up to 2 and 3 fold respectively ^(24)^. The role of animals is controversial, they may either attract sandflies, thus increasing vector density and transmission to humans; or they may be the alternative blood meal source, thereby decreasing transmission ^(25)^. On the other hand a study in India found that keeping animals inside the home is not a risk factor for visceral Leishmaniasis, but improving housing conditions for the poor can decrease its incidence ^(25).^

There was no significant difference in the number of skin lesions between the different age groups, which is similar to previous study in Saudi Arabia ^(26)^, on the other hand we found in our study that the number of lesions is significantly higher in males than females, and higher in farmers compared to other occupations

Less number of lesions was found in patients living near caves, which can be explained that those cases are more aware about the disease. Besides the number was varied across different cities, it was significantly higher in Qalqelia governate.

The duration of the skin lesions was increased in older age groups, less period of time was found in patients who live near caves and who have rocky hyrax and domestic dogs which can be explained that those cases are more aware about the disease and thus earlier presentation. In Salfeet and Tulkarem cases presented to medical attention more lately than other cities.

About one half of the cases were diagnosed during winter and early spring (January till May), the number then decreases and start to rise again in September and October. The seasonal pattern of the disease correlates with the known activity of the vector. There is usually a lag time from the development of skin lesions to the presentation of patients to physicians. This lag in time explains the difference in timing between the peak activity of the sandfly in April and September and the peak of cases in September and October ^(26)^.

The current control strategies for Leishmaniasis depend on reservoir and vector control, active case detection and treatment of their disease and the use of insecticides, more public awareness of prevention and control methods is needed.

### Summary

This study highlights the epidemiology and risk factors of cutaneous leishmaniasis (CL) in the West Bank, Palestine. Our findings indicate a higher prevalence among younger individuals and males, consistent with previous studies in neighboring regions. The predominance of facial lesions and nodules emphasizes the disfiguring nature of CL, underlining its public health impact.

Environmental risk factors, such as proximity to caves and the presence of domestic dogs, appear to modulate disease dynamics. While animals may serve as alternative blood sources for vectors, their role as protective or risk factors remains controversial, necessitating further research.

Geographical disparities in case distribution, with Tubas and Hebron bearing the highest burden, suggest the need for targeted vector control programs in these regions. The seasonal peaks observed align with known sandfly activity patterns, reinforcing the importance of timely interventions.

This study also underscores the socioeconomic dimensions of CL, with farmers and individuals in rural settings facing higher risks.

### Limitations

This study relied on retrospective data, which may have inherent biases.

## Conclusion

Cutaneous leishmaniasis is a significant public health challenge in the West Bank, Palestine.

This study provides critical insights into its epidemiology, clinical features, and environmental risk factors. Effective control measures, including community awareness campaigns, vector control, and improved surveillance systems, are urgently needed.

Future studies should incorporate advanced diagnostic methods and longitudinal designs. particularly in endemic regions like Tubas and Hebron.

## Recommendations

Public health strategies should prioritize education, awareness campaigns, and improved housing conditions to reduce vector-human interactions.

## Data Availability

All data produced in the present study are available upon reasonable request to the authors
All data produced in the present work are contained in the manuscript

## Abbreviations

MOH: Ministry of Health
CI: Cutaneous Leishmaniasis

## Author contributions

EA guarantees the integrity of the entire study. KA and EA participated in conceiving and study design, literature review, supervised data collection, data analysis, manuscript writing. KA, EA and MA performed the material preparation, data collection, and analysis. All authors interpreted the results. EA wrote the first draft of the manuscript, and all authors commented on previous versions of the manuscript. All authors read and approved the final manuscript.

## Acknowledgement

Thank you very much to all the statisticians and health care workers in the Ministry of health surveillance system and preventive medicine department.

